# Transfusion-transmitted infections among blood donors in South-East Italy: Contribution of blood transfusion centres for territory-based surveillance

**DOI:** 10.1101/2023.08.09.23293688

**Authors:** Francescopaolo Antonucci, Antonietta Faleo, Lucia De Feo, Luciano Lombardi, Tommaso Granato

## Abstract

**Background and Objectives:** Screening for Hepatitis B Virus (HBV), Hepatitis C Virus (HCV), Human Immunodeficiency Virus (HIV) infections and Syphilis in blood donors is important to evaluate both risk of transfusion-transmitted infections (TTIs) and their current prevalence in apparently healthy individuals. Apulia (South-East Italy) resulted to be one of the two Italian regions with the highest TTI cases.

Screening tests results of blood donors were analysed, with the purpose to provide updated information on the epidemiology of TTIs.

**Materials and Methods:** 117,454 donors referring to blood transfusion centres of the North Apulia between 2019-2022 were analysed; serum samples underwent analysis for TTIs by chemiluminescent-immunoassay and nucleic-acid-amplification tests.

**Results:** Confirmed reactivities were: 47 HBV (0.04%), 19 HCV (0.02%), 1 HIV (0.001%), and 42 Syphilis (0.04%), respectively. Of 47 HBV-infected donors, 28 were Hepatitis B surface Antigen (HBsAg) positive, including 8 with HBV-DNA positive, and 19 HBsAg negative but with detectable viraemia, identifying the presence of occult B infection (OBI). A higher number of positive cases resulted in the age group 51-65 (34 HBV, 12 HCV, 1 HIV and 25 Syphilis) and over 65 (4 HBV, 5 HCV, 0 HIV and 2 Syphilis) (p-value<0.05). Occasional donors showed higher frequency of TTIs compared to regular donors.

**Conclusion:** The study shows consistent number of undiagnosed TTIs among blood donors, and the importance of transfusion centres for territory-based surveillance that can contribute to the detection of novel TTI cases among asymptomatic people, helping the diagnosis of submerged infectious diseases that are still a global threat.

**HIGHLIGHTS:**

1. Apulia is the second Italian region with the highest number of Transfusion-Transmitted Infections (TTI) among blood donors;
2. In the period 2019-2022, HBV resulted to be the main TTI detected among blood donors, followed by Syphilis, HCV and HIV in North Apulia; the overall TTIs distribution resulted significantly higher among occasional donors than regular donors;
3. Blood transfusion centres may represent important surveillance points for HBV, HCV, HIV and Syphilis and can contribute to the detection of novel TTI cases in apparently healthy individuals.

## INTRODUCTION

Blood and blood products transfusion is a widely used procedure. It is estimated that the use of blood and blood components, applied for different diseases or injuries, is approximately 118 million worldwide every year [1]. Despite this relevant role, blood still may be a source of Transfusion-Transmitted Infections (TTIs); more specifically, the main pathogens that can be transmitted by blood transfusion and that still represent a major public concern [1] are Hepatitis B Virus (HBV), Hepatitis C Virus (HCV), Human Immunodeficiency Virus (HIV) and Treponema Pallidum (etiologic agent of Syphilis).

In the world population, there are approximately 71 million individuals chronically infected with HCV, more than 257 million with HBV, and 37.9 million HIV-infected people [2]. In the European Union, approximately 4.7, 3.9, 6.3 and 1.5 million people cases per year are diagnosed with HBV, HCV, Syphilis and HIV, respectively [3-6], highlighting the importance of further control of the potential novel cases [7]. Based on the reports of World Health Organization (WHO), the prevalence of HBV, HCV, HIV and Syphilis in different parts of the world changes from 0.008% to 6.08% for HBV, 0.004% to 1.96% for HCV, 0.0003% to 7.5% for HIV and 0.0004% to 2% for Syphilis [8-9].

In Italy, based on the Italian surveillance system [10-14], 501 new cases of HBV, 138 of HCV and 5.655 of HIV have been detected in the last four years (2019-2022). Focusing the epidemiological situation in the Apulia Region by considering these data, the number of TTIs detected in general population of this region in 2019, 2020, 2021 and 2022, were respectively: 10, 5, 6 and 7 HBV cases; 2, 1, 3 and 2 HCV cases; 162, 86 and 90 HIV cases (based on our knowledge, no data related with HIV cases in Apulia in 2022 was available when this study was conducted).

Regarding Syphilis, the Italian monitoring system has detected a total of 9,440 new cases (considering both I and II Syphilis) in the period 1991-2020 [15].

Although effective HBV, HCV and HIV treatments have been developing, above all for HCV in the last decade [16-17], considering direct-acting-antivirals for HBV/HCV and antiretroviral therapy for HIV, a great number of infected people remain unaware of their infection [18-19], so most individuals miss diagnosis and treatments and may in turn be an important source of infection transmission.

For all these reasons, in accordance with the WHO recommendations [2], the Italian Ministry of Health Decree of November 2^nd^ 2015 [20] stated that serologic and Nucleic Acid Amplification tests (NAATs) must be performed on all general population attending blood donation with the purpose to discard the infected ones and guarantee transfusion safety [2].

In Italy, all the TTIs detected among blood donors in every region are reported from the involved Blood Transfusion Centres in the National Blood Information System, called SISTRA; this system allows the exchange of information flows between the Italian Ministry of Health and the Regions, favouring interaction between the regional and national level and the timely recording and analysis of data.

Based on SISTRA notifications, the latest surveillance bulletin available from the Italian National Blood Centre [21] reported 1,213 blood donors positive to TTIs in Italy in the year 2021, mainly male and in the age range of 36-65 years old. According to this bulletin, Apulia resulted to be the second region with the higher number of blood donors with TTIs (216 out of 122,844 tested) among all the regions. Of the 216 blood donors with TTI detected in Apulia in 2021, 119 of them were occasional donors tested for the first time (FT donors) for infectious diseases markers and 97 were regular donors repeatedly tested (RT donors) for infectious disease markers, highlighting the higher number of TTIs among the former. More specifically, in terms of number of cases per TTI among FT in Apulia, there were: 52 cases of HBV, 15 cases of HCV, 2 cases of HIV and 50 cases of Syphilis; instead, the number of cases per TTI among RT in Apulia, there were: 73 cases of HBV, 1 case of HCV, 5 cases of HIV and 18 cases of Syphilis.

Considering the higher rate of TTIs in Apulia compared to the other Italian regions [21] in 2021, we analysed in this study the results of screening and confirmation tests of blood donors attending the North Apulia Area (South East Italy) in the period 2019-2022, with the purpose to provide updated information on the epidemiology of TTIs.

## MATERIALS AND METHODS

### Subjects

All blood donors attended one of the seven Blood Transfusion Centres of the North Apulia Area (Foggia, San Giovanni Rotondo, San Severo, Manfredonia, Andria, Barletta and Cerignola) in September 2019-September 2022, were included in this study. All potential donors underwent an initial medical examination; donation permanent exclusion criteria were: malignancies, autoimmune diseases, celiac disease, cardiovascular disease, arterial hypertension, central nervous system diseases, solid organ transplant, coagulopathies, epilepsy, gastrointestinal affections, hepatic affections, haematological affections, immunological affections, renal affections, diabetes, laboratory tests indicating any active infection or past infection related with: HBV, HCV, HIV, Treponema Pallidum, Trypanosoma Cruzi, Coxiella Burnetii. Donation temporary exclusion criteria were: unprotected exposures, intravenous drug use, transfusion or administration of blood components, surgery, dental treatment, tattoos, body piercing, acupuncture, severe allergies, recent vaccinations, drugs administration. Furthermore, the donor was subjected to a check of the haemoglobin value: fundamental information for accessing the donation.

All the information necessary to consider a candidate for blood donation suitable were reported in the pre-donation medical history questionnaire, which was examined by the selecting doctor.

### Microbiological laboratory tests applied for blood donation

Screening tests mandatory for blood donor eligibility were performed in the centralized laboratory of the Transfusion Centre of the University Hospital Riuniti of Foggia.

Two types of screening tests were performed for microbiological analysis of donor blood samples: serologic test and NAAT; the former was based on an enhanced chemiluminescent immunoassay (CLIA) and was performed through VITROS 3600 for the detection of: HCV antibodies (anti-HCV), Hepatitis B surface Antigen (HBsAg), HIV 1/2-P24 Antibody/Antigen (Anti-HIV), and T. pallidum antibodies. Where requested, Hepatitis B core antibody (HBcAb) were assessed as well by VITROS 3600.

As NAAT, a first Transcription-Mediated-Amplification (TMA) was performed through Panther for the detection of viral genome. For samples resulted positive for the first time to TMA, a second TMA was performed for the identification of the viral type (HIV, HCV and/or HBV).

For samples resulting repeatedly positive to one or both the screening tests, confirmatory test by Western-Blotting was performed to confirm the reactivity.

### Data Collection

Blood donor data was collected from Laboratory Information Management Systems (LIMSs) and extrapolated into Excel for the creation of a dataset.

Socio-demographic characteristics of the blood donors and related laboratory test results of HBV, HCV, HIV and Syphilis, were obtained by using the North Apulia Area LIMSs, named WinLab and EmoPuglia. Additional information was collected to distinguish FT donors from RT donors, based on their testing history.

The obtained epidemiological data in this study is related to all donors of the North Apulia Area who confirmed the reactivity to the mandatory tests for the purpose of qualifying blood and blood components [20] and were promptly notified to SISTRA.

### Statistical Analysis

Descriptive statistics were carried considering donors with a confirmed reactivity to HBV, HCV, HIV or Syphilis Antibodies/Antigens. Data analysis was done with the help of Statistical Package for Graph Prism (Version 8.2.1-279). The association of socio-demographic variables, sex and age, with the TTIs was assessed with the Chi-square test (p-value < 0.05 statistically significant). The association of FT and RT donors with the TTIs was assessed with the T-test (p-value < 0.05 statistically significant).

Measurement of the frequency of infection detected at a specified point in time (P, Prevalence) was calculated applying the following formula among the FT donors:

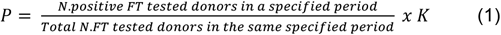

Instead, measurement of new diagnosed cases (I, Incidence) among RT donors were calculated as follows:

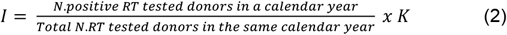

## RESULTS

### Blood donors

The number of blood donors that donated in the North Apulia Area in the period 2019-2022 was 117,454, of which 19,012 FT and 98,442 RT donors. The majority of donors were males (73,996 = 63%). Age distribution was the following: 28,189 (24%) were in the range 18-30, 51,679 (44%) 31-50, 34,062 (29%) 51-65 and 3,524 (3%) over 65 (Table 1). Out of 117,454, 23,213 (19%) donated at the blood transfusion centre of University Hospital Riuniti of Foggia, more specifically: 3,264 (14%) were FT donors and 19,949 (86%) were RT donors, belonging to volunteer associations.

**Table 1:** Demographic characteristics of 117,454 blood donors in the period 2019-2022.

| **Variables** | **Frequency (n)** | **Percentage** |
| --- | --- | --- |
| **Blood donors** |  |  |
| Total | 117,454 | 100 |
| **Age** |  |  |
| 18-30 | 28,189 | 24 |
| 31-50 | 51,679 | 44 |
| 51-65 | 34,062 | 29 |
| >65 | 3,524 | 3 |
| **Sex** |  |  |
| Male | 73,996 | 63 |
| Female | 43,458 | 37 |

### Confirmed TTI cases

Overall, 213 blood donors (0.2%) resulted reactive to laboratory tests for HBV, HCV, HIV, and Syphilis: 56 donors resulted reactive for HBV, 95 HCV, 5 HIV, and 57 Syphilis. Out of 213 reactive cases, 109 confirmed the reactivity and the related TTIs were notified on SISTRA. Of the 109 confirmed TTIs: 47 HBV (0.04%), 19 HCV (0.02%), 1 HIV (0.001%), and 42 Syphilis (0.04%) (Fig. 1A). The distribution of the number of cases per TTI detected in every year is reported in Table 2.

**Table 2:**
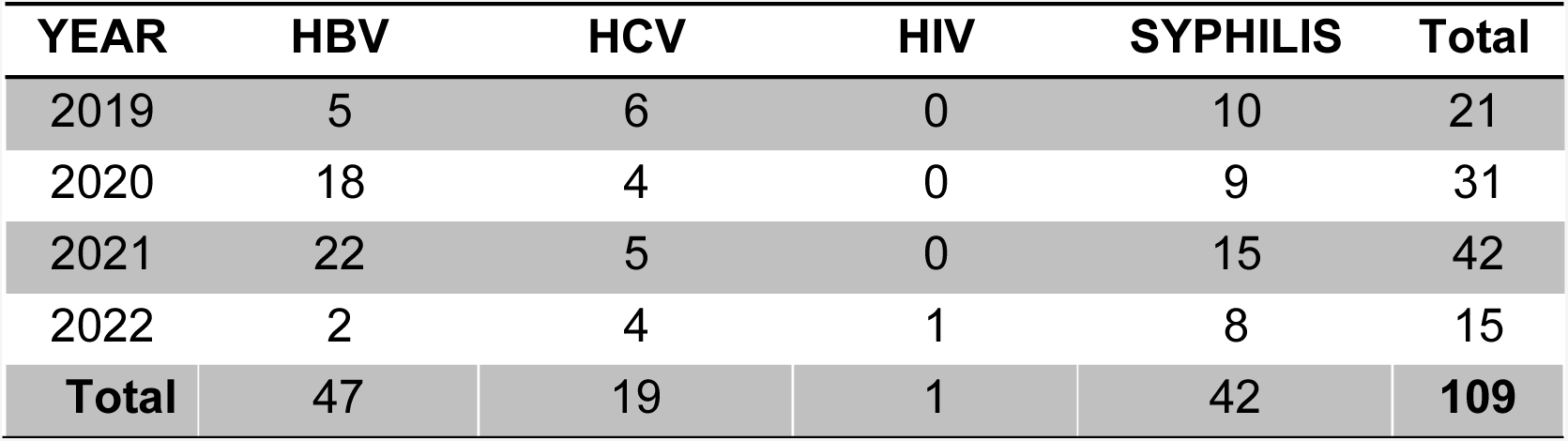
Distribution of TTI cases per year over the period 2019-2022.

**Figure 1:**
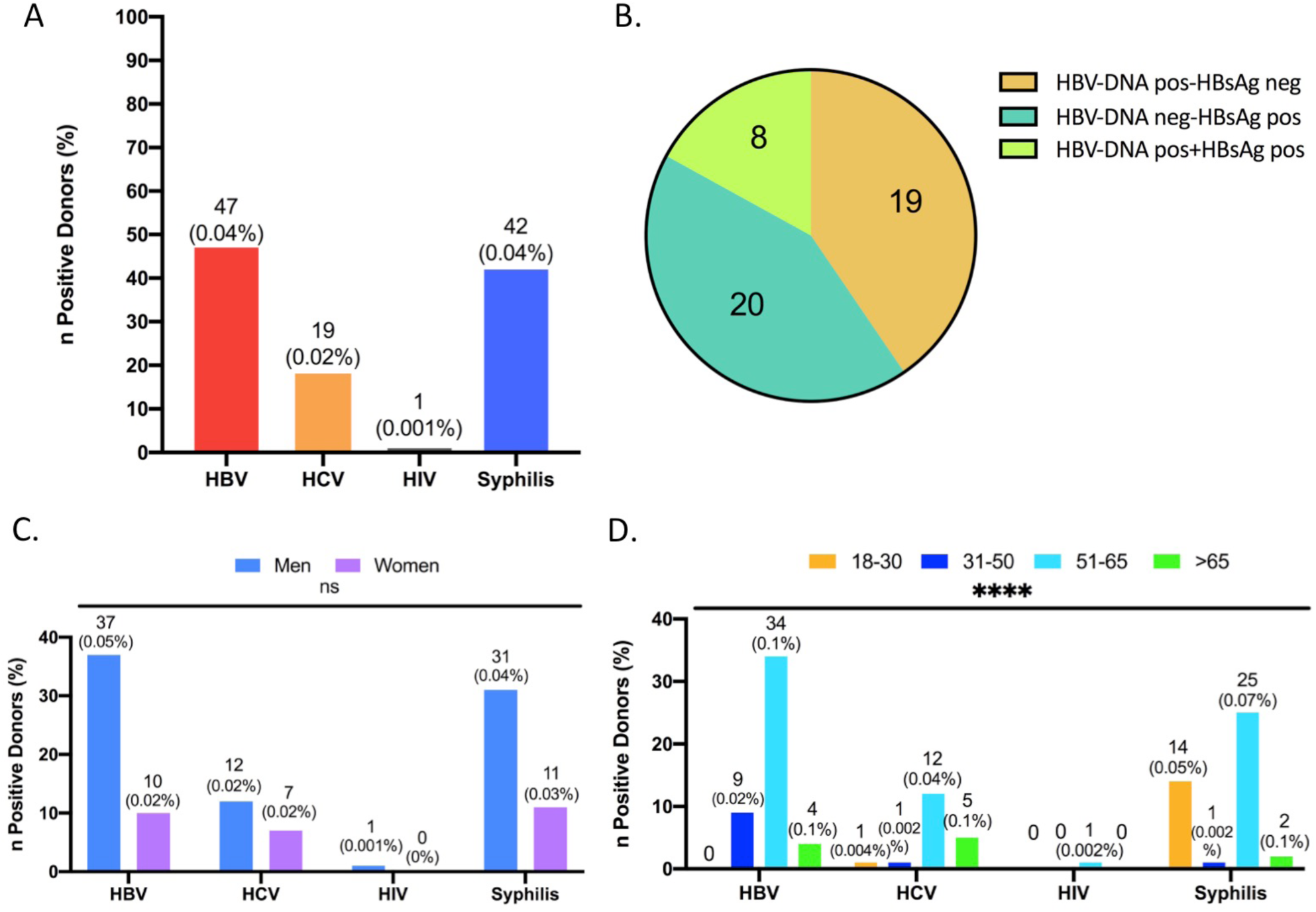
**A**. Number of positive blood donors for HBV, HCV, HIV, and Syphilis. **B**. HBV serological patterns (no. of donors). **C**. Distribution of HBV, HCV and Syphilis cases by sex. The difference in the number of positive cases between men and women is not statistically significant (ns). **D**. Different distribution of HBV, HCV and Syphilis cases by age. Difference among positive cases among age groups is statistically significant (****).

Of the 47 confirmed HBV donors, 28 were HBsAg positive (8 with HBV-DNA positive) and 19 HBsAg negative but with detectable HBV-DNA. Of the latter, fifteen donors were anti-HBc negative and four anti-HBc positive, identifying the presence of seronegative and seropositive occult B infection (OBI) (Fig. 1B).

Nineteen donors were anti-HCV positive with undetectable HCV-RNA. One blood donor was confirmed anti-HIV positive with undetectable HIV-RNA. The difference of TTI cases notified between men (37 HBV, 12 HCV, 1 HIV and 31 Syphilis) and women (10 HBV, 7 HCV, 0 HIV and 11 Syphilis) was statistically not significant (Chi-square, p=0.19), more specifically the percentage of TTIs of men and women were respectively: HBV (0.05% vs 0.02%), HCV (0.02% vs 0.02%), HIV (0.001% vs 0%) and Syphilis (0.04% vs 0.03%) (Fig. 1C).

On the contrary, significant difference (Chi-square, p-value<0.05) was detected among the different age groups. In fact, with the exception of HIV infection, a higher percentage of positive TTI cases resulted in the groups: 51-65 years with 34 (0.1%) HBV, 12 (0.04%) HCV, 1 HIV (0.002%) and 25 (0.07%) Syphilis cases; over 65 with 4 (0.1%) HBV, 5 (0.1%) HCV, 0 (0%) HIV and 2 (0.1%) Syphilis cases. Regarding the other age groups, the positive TTI cases were: 0 (0%) HBV, 1 (0.004%) HCV, 0 (0%) HIV, and 14 (0.05%) Syphilis cases for group 18-30 years; 9 (0.02%) HBV, 1 (0.002%) HCV, 0 (0%) HIV, and 1 (0.002%) Syphilis cases for group 31-50 (Fig. 1D).

Overall 109 confirmed TTIs, 68 were FT donors and 41 were RT donors; the difference of TTIs between FT and RT donors was significant (T-test, p-value<0.05). Prevalence and Incidence of the confirmed TTIs were 131.8 per 100,000 FT donors and 12.4 per 100,000 RT donors, respectively; in particular: 27 HBV cases among FT donors (P=142.0 per 100,000 FT donors), 20 HBV cases among RT donors (I=20.3 per 100,000 RT donors), 17 HCV cases among FT donors (P=89.4 per 100,000 FT donors), 2 HCV cases among RT donors (I=2.0 per 100,000 RT donors), 0 HIV cases among FT donors (P=0 per 100,000 FT donors), 1 HIV cases among RT donors (I=1.0 per 100,000 RT donors), 24 Syphilis cases among FT donors (P=126.2 per 100,000 FT donors) and 18 Syphilis cases among RT donors (I=18.3 per 100,000 RT donors).

## DISCUSSION

Transfusion of blood and blood components is important to save lives and provide a global support to patients. Despite this important aspect, however, transfusion of contaminated blood has a key role in the transmission of blood-borne infectious agents.

It has been estimated the likelihood of TTI in the transfusion of every blood unit is approximately 1% [22]; considering that some of these infections are severe and represents a risk of life because they are incurable or have a difficult treatment process, this estimation is a relatively high rate for transmission of blood–borne diseases. For this reason, TTIs are an important challenge for blood transfusion services and require specific precautions [23]. In the last two decades, vaccination for HBV and improvement on antiviral treatment has greatly reduced the rate of TTIs in different Countries [24], Italy included. The epidemiological control and prophylaxes of the most infectious diseases in Italy have given a great impact on the control of the spread of the related pathogens.

Despite this, there is still a great number of infected people that remain unaware of their infection [18-19]; this will cause a missed diagnosis in these individuals, missed treatments and increased risk of infection transmission from this source.

In this study, we analised the number of TTIs related with HBV, HCV, HIV and Syphilis in a large number of blood donors attending the Blood Transfusion Centres of the North Apulia Area over the period 2019-2022.

Based on our results, considering the TTIs in the last year 2022, the highest number of TTIs cases among blood donors in the North Apulia Area is Syphilis (8 cases), followed by HCV (4 cases), HBV (2 cases) and HIV (1 cases). Despite this increase of Syphilis cases in the last year observed in this Area, HBV represented the main TTI detected among the blood donors in the last 4 years period (2019-2022), with a distribution of confirmed cases per year of 5 in 2019, 18 in 2020, 22 in 2021, and 2 in 2022.

High cases of chronic hepatitis viruses HCV, together with HBV, is more frequent in blood donors compared to those detected in the general population, which mainly reports acute infections [21]. Furthermore, the highest risk of TTI related with HBV might be attributed to the interval between initial HBV infection and the detection of HBsAg, resulting in a long window phase during which the virus is transmissible, making this pathogen more difficult to be diagnosed.

Overall, considering the total number of detected TTIs among 117,454 blood donors attending blood transfusion centres of the North Apulia Area in the last four years, the majority of blood donors resulted positive to TTIs were men (81 men vs 28 women), although the different distribution was statistically not significant (p-value>0.05), due to the higher number of men involved in the donation compared to women [25].

Differently, the distribution of TTIs resulted to be significantly different (p-value<0.05) among the age groups considered in this study; more specifically, higher number of TTIs related with HBV, HCV, HIV and Syphilis was observed in the over 51 age group. HBV was the most frequent TTI detected among blood donors of the 31-50 age group; similarly, Syphilis was the most frequent TTI detected among blood donors of the 18-30 age group. Another aspect noticed in our study is related with the different distribution of TTIs between the two blood donors categories attending the blood transfusion centres of the North Apulia Region: FT donors and RT donors. For the instance, the number of TTIs detected among FT donors was significantly higher (p-value<0.05) than the TTIs detected in RT donors; according to our study, HBV and Syphilis were the most common infections detected with higher percentage among FT donors compared to RT donors.

Interestingly, all the infected donors were unaware of their status, and this highlights the importance of territory-based surveillance to both keep blood donation safe and monitor the main infectious diseases that can spread from asymptomatic people.

A limitation of the study is its retrospective nature, which cannot provide information on risk factors and disease status.

In conclusion, blood transfusion centres, supported by the contribution of associations that recruit volunteer regular donors through blood donation awareness campaigns, represent important surveillance points of a large number of people for HBV, HCV, HIV and Syphilis and can contribute to the detection of novel cases in apparently healthy individuals and favour the emergence of submerged cases of infectious diseases.

## Data Availability

All data produced in the present study are available upon reasonable request to the authors

## ACKNOWLEDGEMENTS

T.G., L.L. and A.F. performed medical examination of blood donors; F.A. and L.D. assessed virological and serological laboratory results; F.A. acquired data, analysed the data, wrote the manuscript, reviewed it and edited it; T.G. checked the manuscript.

